# PRACTICE AND ASSOCIATED FACTORS OF PEDIATRICS EMERGENCY TRIAGE AMONG HEALTHCARE PROVIDERS WORKING AT TERTIARY HOSPITALS IN WEST OROMIA, ETHIOPIA, 2025

**DOI:** 10.1101/2025.06.17.25329821

**Authors:** Gurmessa Dessale, Teshale Mulatu, Amanuel Tesfaye, Dereje Temasgen, Duguma Debela

**Affiliations:** School of Nursing, College of Health Sciences, Wallaga Unversity, Nekemte, Ethiopia; School of Nursing, College of Health Sciences, Assosa University, Assosa, Ethiopia; School of midwifery, College of Health Science, Wallaga University, Nekemte, Ethiopia; Higher education and training officer, Rift Valley University head office, Addis Ababa, Ethiopia

**Keywords:** Practice, Pediatric emergency triage, Ethiopia

## Abstract

**Background:** The mortality rate in pediatric emergency rooms within developing countries remains alarmingly high, primarily due to preventable and treatable conditions. effective pediatric emergency triage is critical for identifying and prioritizing children with serious health issues. However, there is currently limited evidence regarding the practices and factors influencing pediatric emergency triage among healthcare providers, specifically in Western Oromia, Ethiopia.

**Methods:** An institutional-based cross-sectional study was conducted from February 30 to March 30, 2025 on a total of 422 health care providers selected using a simple random sampling technique. The data were collected using pretested and interviewed based questionnaires. The data were cleaned, entered into Epi-data 4.7.0.0 and exported to SPSS-27 for analysis. Bivariate and multivariate logistic regressions were done to identify factors associated with triage practice. Variables with a p-value of ≤ 0.05 at 95% CI were considered statistically significant variables.

**Results:** From a total of 422 respondents, 414 (98.10 %) of them gave complete responses. Majority of respondents (60.1%) had a good triage practice. Variables like training on guideline [AOR = 1.74, 95% CI: (1.07 - 2.84)], good knowledge [AOR = 3.40, 95% CI: (2.07 - 5.57)], triage experience [AOR = 2.76; 95% CI:(1.66 - 4.58)], presence of essential drugs and equipment [AOR = 2.16; 95% CI: (1.30 - 3.59)], and good attitudes [AOR = 3.26; 95% CI: (1.96 - 5.42)], were significantly associated with triage practice of healthcare providers.

**Conclusion and Recommendation:** About three fifth of respondents had a good triage practice. Key factors associated with triage practice included good knowledge, presence of essential drugs and laboratory support, triage experience, training on guidelines, and attitude of healthcare providers were factors associated with triage practice. Therefore, stakeholders including hospital managers should implement targeted strategies like training on increasing knowledge and enhancing positive attitude among healthcare providers to prevent the occurrence of poor triage practice.

## Introduction

A pediatric emergency is a serious place that affects newborns, infants, or teenagers, requiring immediate medical attention to prevent significant harm or death (1). Triage is the rapid assessment, categorization, and allocation of patients to determine the urgency of their need for medical care, particularly in the emergency department (2). Pediatric emergency triage is the process of quickly evaluating and categorizing children when they first present to an emergency department to single out those suffering from serious conditions among all initially presenting complaints (3). Globally, pediatric emergency triage is an essential procedure in healthcare systems, ensuring that children receive prompt and appropriate medical care based on the severity of their illnesses (4). Pediatric emergency triage involves quickly evaluating and categorizing children to prioritize those with serious conditions (5).

The WHO framework for ETAT emphasizes early detection and timely interventions to reduce morbidity and mortality, adapting advanced pediatric life support guidelines for developing countries (1). Worldwide, PET protocols are integral to healthcare systems (4,5). Adaptations of international guidelines like the Emergency Triage Assessment and Treatment protocols within Ethiopia reveal, provide valuable structure, however their successful implementation heavily depends on factors including provider competency to practice it. The Triage was introduced in Ethiopia in 2014, designed to be performed within 20 seconds of a child’s arrival (1,6).

Robust pediatric emergency triage systems are important as they help manage high patient volumes, ensure rapid decision-making, and optimize the allocation of scarce yet essential resources. In emergencies such as mass casualties and disasters, effective triaging is essential for prioritizing critical cases and efficiently allocating resources to achieve optimal outcomes (7). The WHO has proposed guidelines, particularly for low-income settings, to establish comprehensive pediatric emergency triage protocols addressing areas such as hypoxemia detection, fluid management, seizure management, and others (8). However, under-triage and over-triage scenarios can occur. Under-triage happens when patients with rapidly deteriorating conditions are not identified and missed, while over-triage occurs when patients with acute but non-life-threatening illnesses are prioritized, resulting in a waste of medical resources (9).

Healthcare providers in emergency departments must quickly distinguish between patients requiring immediate treatment and those whose conditions allow for delayed assessment, ensuring optimal patient care under high clinical demands (9). The WHO recommends the application of triage at all levels of the facility. However, its implementation triage is limited to tertiary hospital in our country due different reason. Therefore this study was conducted at the only tertiary hospital in West Oromia (1). By evaluating the outcomes of these tertiary hospitals, we can generate evidence to support the expansion of effective triage systems to secondary hospitals, health centers, and non-governmental hospitals.

In these institutions, healthcare providers (doctors and nurse) are at the frontline, responsible for implementing triage protocols that can literally mean the difference between life and death. However the inconsistent application of triage guidelines and varied pediatric presentations underscore the need to thoroughly explore pediatric emergency triage practices and their influencing factors. Therefore this study aims to study the practices of healthcare providers and their influencing factors by including socio-demographic, Institutional-related and healthcare worker-related factors aspects.

## Materials and Methods

This study was conducted in the pediatric emergency departments of selected tertiary hospitals in West Oromia, Ethiopia. This West Oromia includes six tertiary hospitals such as Ambo University Referral Hospital, Dambi Dollo University Comprehensive Specialized Hospital, Jimma University Medical Center, Nekemte Comprehensive Specialized Hospital, Wallaga University Comprehensive Specialized Hospital, and Mettu Karl Comprehensive Specialized Hospital. Ambo University Referral Hospital, located in Ambo, East Showa, is 114 km from Addis Ababa and serves a population of approximately 5 million, functioning as a teaching hospital that offers multidisciplinary services.

Wallaga University Comprehensive Specialized Hospital, also in Nekemte and approximately 328 km from Addis Ababa, provides comprehensive multidisciplinary services for numerous patients in the East and West Wallaga zones. Jimma Medical Center, located 350 km from Addis Ababa in Jimma, serves over 20 million people from surrounding regions, delivering extensive multidisciplinary care annually. Initially, the study collected information on triage services in various government and private health facilities, with a specific focus on pediatric emergency triage of facility in West Oromia. Then, the findings indicated that general hospitals, referral hospitals, primary hospitals, and health centers do not have dedicated pediatric emergency triage services. Instead, these facilities implemented a central triage system that integrates both adult and pediatric triage service. However, pediatric emergency triage services are specifically provided in tertiary hospitals.

There are six tertiary hospitals. From these, this study focused on three tertiary hospitals in West Oromia, selected through simple random sampling. the lottery method was employed, often involving drawing from a container. While, Nekemte Comprehensive Specialized Hospital was utilized for a pretest, a total of 1,298 healthcare providers work across all included tertiary hospitals. The providers are distributed as follows: AURH (283), NCSH (141), DDUCSH (115), WUCSH (182), MKCSH (165), and JUMC (412) from HMIS of each hospital. The study was collected data from three tertiary hospitals: JUMC (412), AURH (283), and WUCSH (182). In total, 877 healthcare providers participated in the study. The study was conducted from February 30 to March 30, 2025.

### Study design and Population

An institutional-based cross-sectional study was employed. All healthcare providers working in various units of selected tertiary hospitals in western Oromia were included in the study. All healthcare providers who provided free services in the unit, those HCPs who do not rotate to the pediatric emergency triage unit, and healthcare providers with less than six months of experience were excluded.

### Sample size determination and sampling procedures

The sample size was determined using a single population proportion formula with the following assumptions: Z= the standard average deviation at 95% confidence level; =1.96, d= margin of error that can be tolerated, 5% (0.05), and P is the proportion of triage practice skills at Addis Ababa study (49.7%).

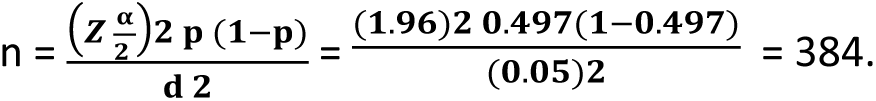

The calculated sample size was 384. By adding a 10% non-response rate, the final sample size was 422. A systematic sampling technique was employed to pick healthcare providers from each hospital after samples were proportionally allocated for all tertiary hospitals. First list of healthcare providers working PET in selected tertiary hospitals were taken from their HMIS of hospitals. The total number of healthcare providers in these three selected tertiary hospitals is 877. Then, the number of healthcare providers to include from each hospital was determined using the formula: ni = n/N × Ni, where N is the total number of healthcare providers in the three tertiary hospitals(877), Ni is the sample size (422), and n is the total number of healthcare providers in each hospital. Each allocated healthcare provider was assigned a unique identification code. These codes were entered into SPSS (Version 27) for randomized selection. These codes were entered into SPSS (Version 27) for randomized selection, resulting in the selection of 198 healthcare providers from JUMC, 136 from AURH, and 88 from WUCSH, culminating in a total sample size of 422. The generated random numbers/code were printed and subsequently used to identify the corresponding healthcare providers within each hospital’s records. A simple random sampling method selected by using lottery methods was then employed to select specific healthcare providers from each hospital’s list. The data collector obtained informed consent from each selected provider before collecting the necessary data.

### Study Variables

#### Dependent variable

Practice of pediatric emergency triage

#### Independent variable

Socio-demographic characters: age, sex, marital status, level of education and work unit. Healthcare provider-related factors: training on ETAT, knowledge of HCP, read guideline, type of profession, work experience, triage experience, attitude of HCP with ETAT.

Institutional-related factors: presence of management support, presence of standardized guideline and protocols, and presence of essential drugs and laboratory support.

#### Data collection tools and procedures

The data collection tool/instrument was adapted from various literatures of a prior study conducted and contextualized to the study setting. The tool was prepared and administered in the English language. The data collection tool consists of 63 questions categorized into three sections that gather information regarding practice and associated factors including socio-demographic, healthcare providers and institutional related factors.

Section 1: determinant factors about pediatrics emergency triage among healthcare providers. Part 1: The socio-demographic questionnaire consisted of 11 questions. Part 2: healthcare provider’s related factors on pediatrics emergency triage consisted of 54 with sub-items questions, including three multiple-choice formats and a set of True/false/I doesn’t know/ questions. The participants responses a correct were scored as 1 (or marked as “True”), while incorrect responses were scored as 0 (or marked as “No” and “I don’t know”). Additionally, ten attitude-related questions prompted participants to express their opinions on healthcare providers’ pediatric emergency triage practice skills, using a scale from 1 (strongly disagree) to 5 (strongly agree). A participants scoring above the mean on attitude questions received a score of 1, while those scoring below received a 0. Scores were assigned to responses of “Strongly Disagree” (1 point) and “Strongly Agree” (5 points), with reverse coding applied for negatively phrased questions to accurately reflect participants’ attitudes.

Section 2: Practice/skills of respondents towards triage were assessed by using 27 questions divided into three categories: quick assessment, patient classification, and patient distribution. Respondents were given a scale with a 5-point Likert-type scale for their measurements, with ratings from “5 = Always, 4 = Often, 3 = sometimes, 2 = seldom, 1 = rarely”, and computed revisable for negative statements of items. The sum score was classified into 2 levels (good practice and poor practice). Section 3: institutional-related factors have two part; part 1: Responses to each question are given as either “yes” or “no.” A “yes” response is assigned a score of 1, while a “no” response is assigned a score of 0. Part 2: This section presents a descriptive analysis of institutional factors influencing pediatric emergency triage practice, identifying both barriers to effective triage and enablers of optimal triage.

An interviewed-administered based questionnaire was utilized to gather data from study respondents. Actual data collection was carried out by three BSc nurses with prior experience in data collection, while three MSc nurses were recruited as supervisors. Data facilitators carefully filled and ensuring the questionnaires completed all items accurately. Finally, the data facilitators collected the filled data after checking for the completeness of the data.

#### Operational Definition and Definition of Terms

**Triage practice:** It is refers to the systematic methodology employed by healthcare providers to rapidly assess, classify, and prioritize patients based on clinical urgency in pediatric emergency settings (10). Practice triage was evaluated using a standardized scoring system where: Good practice: those participants scoring ≥ 3.34 (the mean score) on correctly answered practice questions and Poor practice: Participants scoring below the mean value of 3.34 on practice questions were categorized as having poor practice.

**Triage knowledge:** it is refers the critical understanding of facts and procedures that healthcare professionals need in pediatric emergency triage to perform evaluations, categorize patients, and assign them to the appropriate care. This knowledge is assessed based on cut off mean scores, where a mean score below 1.82 indicates a poor knowledge toward triage practice, whereas a mean score above or equal 1.82 reflects a good knowledge.

**Triage attitude:** it refers to healthcare providers’ perceptions and beliefs regarding pediatric emergency triage, measured through responses to standardized attitude questions. Responses were recorded using a five-point Likert scale (1 = strongly disagree to 5 = strongly agree) and categorized based on composite mean scores. Good attitude: Those participants who scored ≥ 3.04 (mean value on attitude questions) and Poor attitude: Those participants who scored < 3.04 on attitude questions.

**Healthcare providers:** In this study, the term refers to both doctors and nurses currently working in tertiary hospitals in West Oromia.

### Data Quality Assurance

Data quality was assured through a one-day orientation for data facilitators and the supervisor, as well as pretesting the data collection tool prior to the main study. The orientation covered data collection techniques, the purpose of data collection, the content of the questionnaires, methods for approaching respondents, and how to resolve any issues that may arise during the data collection process. Throughout the data collection period, the principal investigator and supervisor provided daily oversight to maintain the completeness and quality of the data. Upon completion of the data collection, each questionnaire was carefully reviewed for completeness, and data cleaning was done involve checking for inconsistencies, numerical errors, and missing parameters.

A pretest was conducted at Nekemte Comprehensive Specialized Hospital one week prior to the actual data collection to assess the consistency and clarity of the language used in the tool. The validity of the tool was assessed by a jury of two experts in the field of emergency, and their recommended modifications had been done. The pretest was conducted by takng 5% (21 respondents ) of the total sample size and administering the questionnaire to them. Following the pretest, an evaluation of the language clarity of the tool was performed, and necessary modifications were implemented. Additionally, inter-item consistency of the tool was assessed using Cronbach’s coefficient alpha and with each subscale to test internal consistency. The results showed a coefficient of 0.82 for practice, 0.97 for Healthcare providers, and 0.91 for attitude in relation to emergency triage practice.

### Data Processing and Analysis

After the data collection, the data were rechecked for completeness, cleaned, coded, and entered into Epidata version 4.7.0.0, and subsequently exported to SPSS version 27 for analysis. Appropriate coding was done at each step for the variables. The missing data were handled by imputation (giving code), which involved replacing the missing data with certain values. Descriptive statistics, including frequency, percentages, mean, and standard deviation, were carried to summarize the data. Then, binary logistic regression was conducted to evaluate the unadjusted relationship between the independent variables and the dependent variable. All variables with a p-value of less than 0.25 were considered candidates for multivariate logistic regression to control for possible confounding effects. Then, to assess the independent effects of each variable on the outcome, a multivariate logistic regression analysis was conducted. The model’s goodness-of-fit was checked by using the Hosmer-Lemeshow test, which indicated adequate fit with a non-significant p-value of 0.37, suggesting the model appropriately represented the observed data. Multicollinearity was checked using the variance inflation factor (VIF) and its values lie between 1.038 and 2.048, which is interpreted as no multicollinearity among the predictor variables. Then, variables with p-value ≤ 0.05 were declared statistically significant associated factors with practice. Finally, the analysis result was presented in texts, tables, and graphs as appropriate.

## RESULT

### Socio-demographic characteristics of nurses

From the total of 422 study participants, 414 were included in the study; resulting in a response rate of 98.10% (eight study participants were omitted from the analysis due to incomplete data from two respondents, one refused, and five unavailable respondents). More than half 218 (52.7%) of the participants were male. The average age of respondents was 35 ± 6.47 (SD) years, and 247 (59.7%) of the respondents were below the age of 35 years. More than two third of the participants 281 (67.9 %) were married.

Regarding level of education status more than two fifth, 195 (47.1%) of healthcare providers held a bachelor’s degree. lastly, concerning working units, 116 (28.0%) participants worked in the ward unit (Table 2).

**Table 1:**
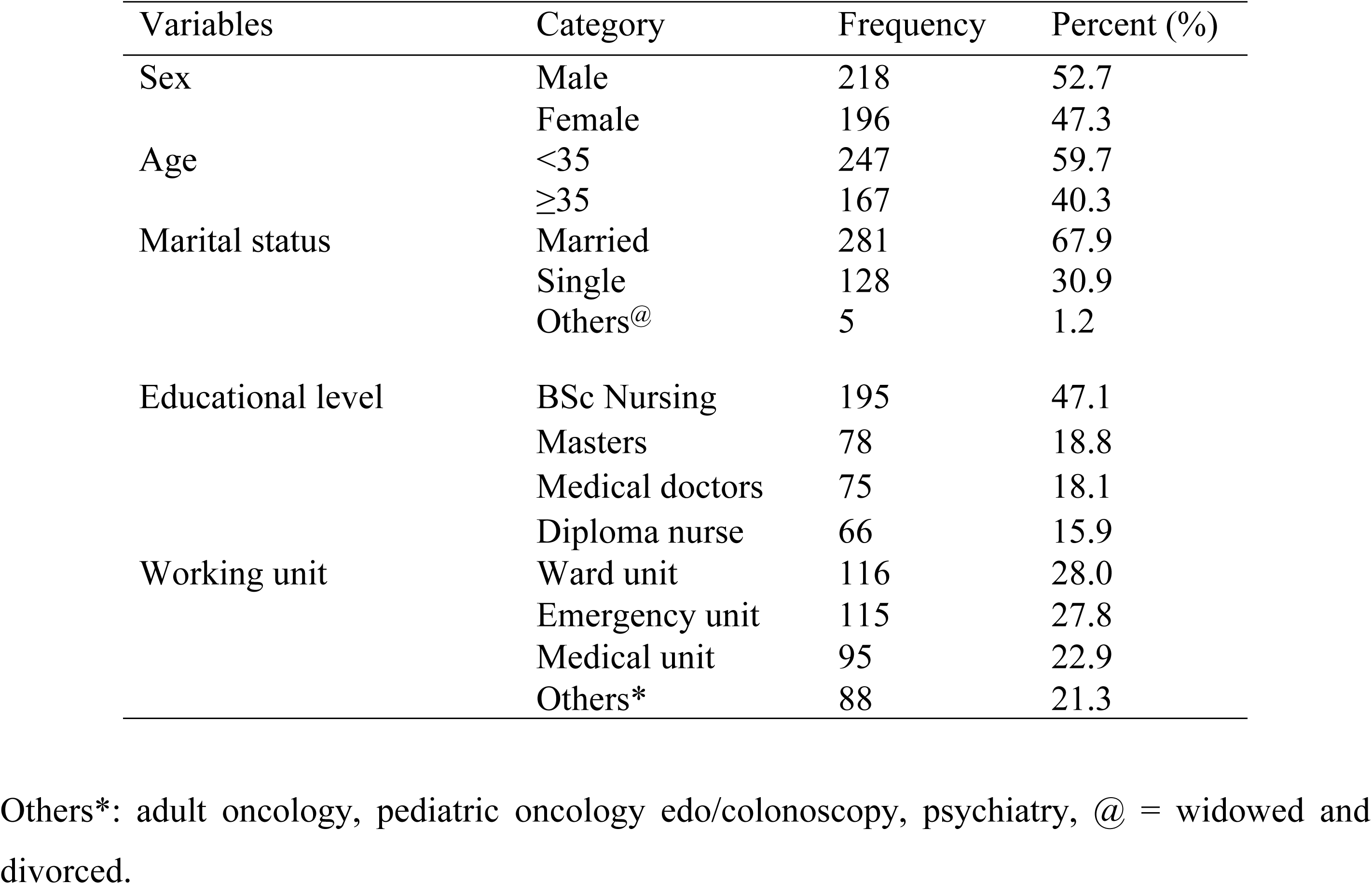
Socio-demographic characteristics of healthcare providers working in Tertiary Hospital, West Oromia, Ethiopia, 2024 (n=414).

**Table 2:** Triage practice subscale of patient categorization and patient allocation among healthcare providers working in tertiary hospitals, West Oromia, Ethiopia, 2025 (n=414).

| Variable | | Mean ± SD | Scoring level | |
| --- | --- | --- | --- | --- |
|  |  |  | Good | Poor |
|  |  |  | N (%) | N (%) |
| Subscale | Patient Assessment | $3.28 \pm 0.67$ | 243 (58.7) | 171 (41.3) |
| | Patient categorization | $3.50 \pm 0.79$ | 264 (63.8) | 150 (36.2) |
| | Patient Allocation | $3.48 \pm 0.80$ | 275 (66.4) | 139 (33.6) |

SD = Standard deviation. \* = Five-point Likert scale measuring practice

### Level of triage practice

The study revealed, of total respondents mean scores were classified as Good (≥ 3.34) and Poor (< 3.34). Among the healthcare providers assessed, 249 (60.1%) demonstrated good practice, while 165 (39.9%) exhibited poor practice in pediatric emergency triage (Figure 4).

The study revealed the overall mean ± SD score for patient assessment practices in pediatric emergency triage was 3.28 ± 0.67, with 243 (58.7%) healthcare providers demonstrating proper patient assessment. Similarly, the mean ± SD score for patient categorization practice of pediatric emergency triage among healthcare providers was 3.50 ± 0.79, indicating a good level of practice. Specifically, 264 (63.8%) of respondents reported good patient categorization practices. Lastly, the mean ± SD score for patient allocation practice of pediatric emergency triage was 3.48 ± 0.80. Among all respondents, 275 (66.4%) of healthcare providers properly practice patient allocation (Table 3).

**Table 3:**
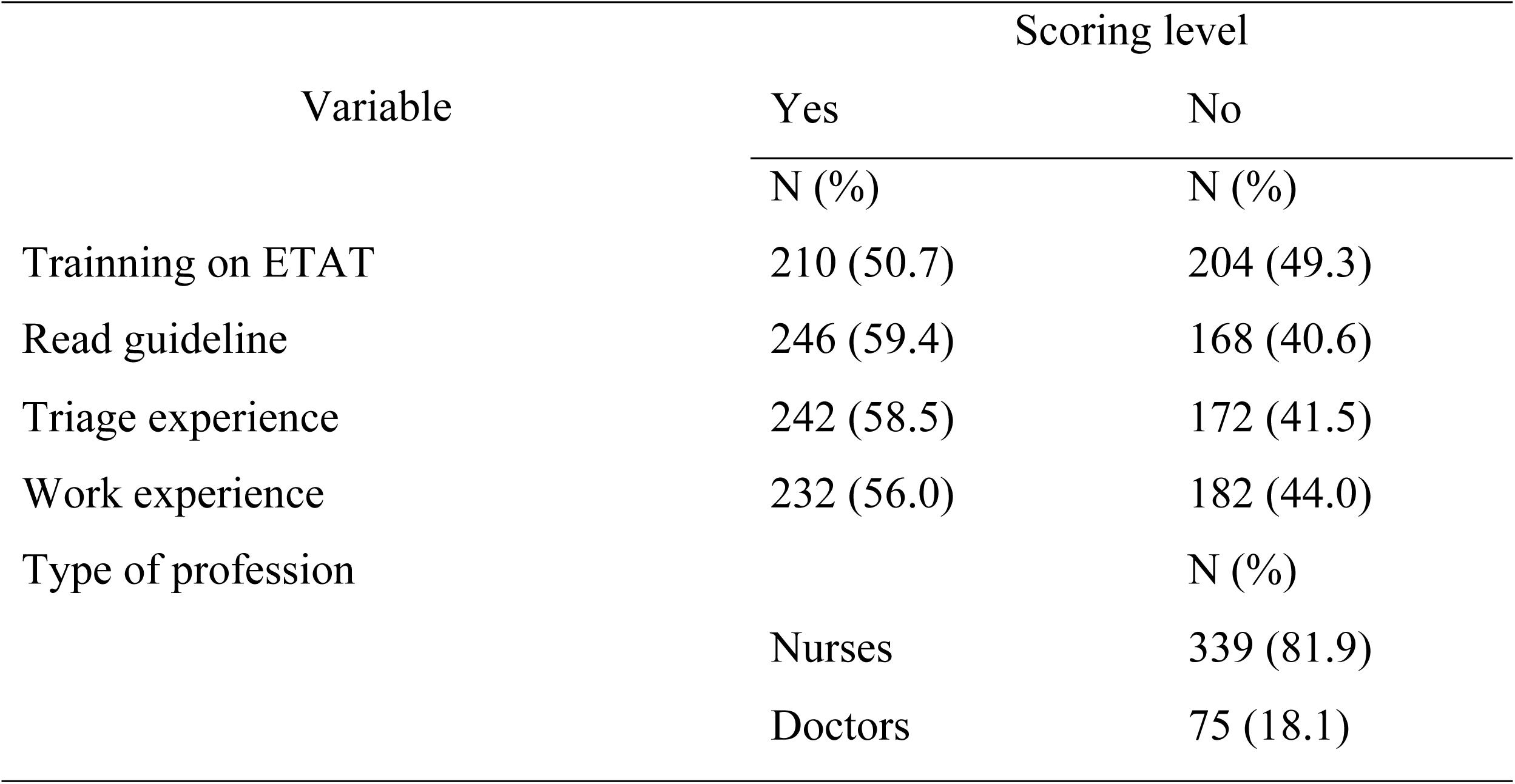
Responses of healthcare providers regarding training, read, profession ,triage and work experience at a selected Tertiary Hospitals in West Oromia, Ethiopia, in 2025 (n = 414).

### Healthcare provider related factors

The study revealed found that more than half the participants 242 (58.5%) reported having prior triage experience in pediatric emergency care. Additionally, over half of the respondents, 232 (56.0%) had fewer than five years of work experience, while 182 (44.0%) had more than five years of experience. Among all participants, 210 (50.7%) indicated that they had received training on ETAT guideline. Likewise, More than half 246 (59.4%) of Healthcare providers was reading guidelines related to pediatric emergency triage guidelines and more than three quarters of the participants, 339 (81.9%) were identified as Nurses (Table 4).

**Table 4:** Knowledge of healthcare providers regarding important priority signs for triage and assessment in pediatric emergency triage at a selected Tertiary Hospital in West Oromia, Ethiopia, in 2025 (n = 414).

| Priority sign | Response | | | Mean ± SD |
| --- | --- | --- | --- | --- |
|  | True | False | I don't |  |
| Tiny baby | 180 (43.5) | 108 (26.1) | 126 (30.4) | $1.87 \pm 0.85$ |
| Temperature | 214 (51.7) | 85 (20.5) | 115 (27.8) | $1.76 \pm 0.86$ |
| Trauma | 162 (34.1) | 141 (34.1) | 111 (26.8) | $1.88 \pm 0.80$ |
| Pallor | 189 (45.7) | 122 (29.5) | 103 (24.9) | $1.79 \pm 0.82$ |
| Poisoning | 183 (44.2) | 99 (23.9) | 132 (31.9) | $1.88 \pm 0.87$ |
| Pain | 187 (45.2) | 116 (28.0) | 111 (26.8) | $1.82 \pm 0.83$ |
| Respiratory distress | 201 (48.6) | 106 (25.6) | 107 (25.8) | $1.77 \pm 0.83$ |
| Restless | 201 (48.6) | 108 (26.1) | 105 (25.4) | $1.77 \pm 0.82$ |
| Referral | 195 (47.1) | 94 (22.7) | 125 (30.2) | $1.83 \pm 0.86$ |
| Malnutrition | 199 (48.1) | 103 (24.9) | 112 (27.1) | $1.79 \pm 0.84$ |
| Oedema | 211 (51.0) | 97 (23.4) | 106 (25.6) | $1.75 \pm 0.84$ |
| Burns | 200 (48.3) | 96 (23.2) | 118 (28.5) | $1.80 \pm 0.86$ |

### Knowledge of health care providers

This study showed that the overall mean ± SD score for healthcare provider’s knowledge was 1.82 ± 0.50 and 58.0% of healthcare providers had good knowledge (Figure 5).

Knowledge of healthcare providers regarding important priority sign.

The result of the study shows that the mean scores for knowledge of important priority signs indicators of knowledge HCPs were 1.81 ± 0.56. Based on these scores, 56.8% of HCPs had good knowledge of important priority signs, while 43.2% of respondents had poor knowledge.

The study revealed significant variability in healthcare providers’ ability to recognize important priority signs of pediatric triage indicators. Conditions such as temperature 214 (51.7%) and edema 211 (51.0%) were correctly identified by over half of the respondents. However, gaps in knowledge were evident, as 25-30% of providers either responded incorrectly or were unsure about priority signs (Table 5).

**Table 5:** The knowledge of healthcare providers concerning immediate sign for triage and assessment in pediatric emergency triage at a selected tertiary hospital in West Oromia, Ethiopia, in 2025 (n = 414).

| Emergency sign | Response | | | Mean± SD |
| --- | --- | --- | --- | --- |
|  | True | False | I don't |  |
| severe respiratory distress | 224 (54.1) | 94 (22.7) | 96 (23.2) | 1.69 ± 0.82 |
| Obstructed or absent breathing | 225 (54.3) | 99 (23.9) | 90 (21.7) | 1.67 ± 0.81 |
| Circulation problem/signs of shock | 218 (52.7) | 93 (22.5) | 103 (24.9) | 1.72 ± 0.84 |
| Coma or Unconscious | 161 (38.9) | 109 (26.3) | 144 (34.8) | 1.96 ± 0.86 |
| Convulsion | 175 (42.3) | 125 (30.2) | 114 (27.5) | 1.85 ± 0.82 |
| Severe dehydration | 178 (43.0) | 112 (27.1) | 124 (30.0) | 1.87 ± 0.85 |
| Bleeding child | 191 (46.1) | 104 (25.1) | 119 (28.7) | 1.83 ± 0.85 |
| Immediate poisoning | 175 (42.3) | 127 (30.7) | 112 (27.1) | 1.85 ± 0.80 |
| Major trauma | 190 (45.9) | 95 (22.9) | 129 (31.2) | 1.85 ± 0.87 |
| Disabilities | 193 (46.6) | 110 (26.6) | 111 (26.8) | 1.80 ± 0.84 |
| Central cyanosis | 185 (44.7) | 120 (29.0) | 109 (26.3) | 1.82 ± 0.82 |

SD = Standard deviation. \* = three-point Likert scale measuring emergency sign indicator

### Knowledge of healthcare providers regarding important immediate signs

The result of the study shows that the mean scores for knowledge of important immediate signs indicators subscale knowledge HCPs were 1.81 ± 0.54. Of the 414 respondents, 248 (59.9%) had good knowledge, while 166 (40.1%) exhibited poor knowledge of essential emergency signs.

The analyses of individual responses items revealed that severe respiratory distress 224 (54.1%), obstructed breathing 225 (54.3%), and circulatory shock 218 (52.7%) were correctly identified by over half of by most respondents (Table 6).

**Table 6:** Knowledge of Healthcare providers regarding essential drug needs in pediatric emergency triage at a Selected Tertiary Hospital in West Oromia, Ethiopia, 2025 (n = 414).

| Essential drugs | Response | | | Mean ± SD |
| --- | --- | --- | --- | --- |
|  | True<br>N (%) | False<br>N (%) | I don't<br>N (%) |  |
| Half-strength Darrow's with 5% glucose | 204 (49.3) | 96 (23.2) | 114 (27.5) | $1.78 \pm 0.85$ |
| Ringer's lactate or 0.9% normal saline | 189 (45.7) | 104 (25.1) | 121 (29.2) | $1.84 \pm 0.85$ |
| 40% glucose | 184 (44.4) | 122 (29.5) | 108 (26.1) | $1.82 \pm 0.82$ |
| IV Diazepam or Lorazepam | 179 (43.2) | 110 (26.6) | 125 (30.3) | $1.87 \pm 0.85$ |
| Adrenaline Iv | 171 (41.3) | 114 (27.5) | 129 (31.2) | $1.90 \pm 0.85$ |
| Salbutamol Puff | 181 (43.7) | 119 (28.7) | 114 (27.5) | $1.84 \pm 0.83$ |

SD = Standard deviation. DW= Dextrose in water, IV= Intravenous.\* = three-point Likert scale measuring essential drugs indicator

### The knowledge of HCP toward essential drugs

This result showed that the HCPS knowledge mean score for essential drugs was 1.84 ± 0.57. of 215 providers (51.9%) had good knowledge while, 199 (48.1%) showed poor knowledge of essential drugs.

The study revealed moderate but concerning gaps in healthcare providers’ knowledge of essential pediatric emergency drugs, with correct identification rates ranging from 41.3% (Adrenaline Iv) to 49.3% (half-strength Darrow’s solution). All essential medications showed similar patterns of knowledge deficiencies, with 26.1-31.2% of providers expressing uncertainty and 23.2-29.5% providing incorrect responses (Table 7).

**Table 7:**
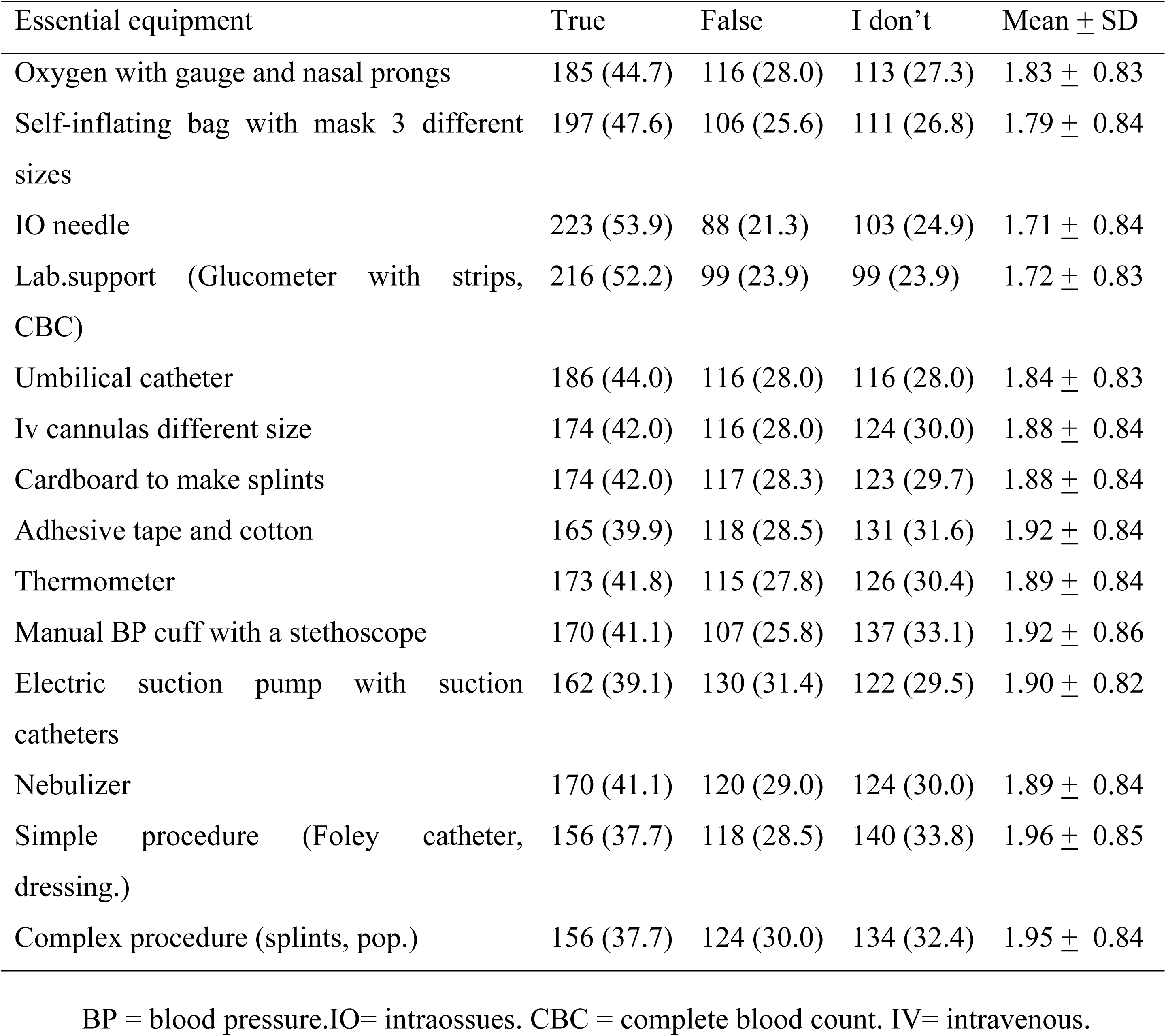
Knowledge of healthcare providers regarding essential equipment needs in pediatric emergency triage at a selected tertiary hospital in West Oromia, Ethiopia, 2025 (n = 414).

| Essential equipment | True | False | I don't | Mean ± SD |
| --- | --- | --- | --- | --- |
| Oxygen with gauge and nasal prongs | 185 (44.7) | 116 (28.0) | 113 (27.3) | 1.83 ± 0.83 |
| Self-inflating bag with mask 3 different sizes | 197 (47.6) | 106 (25.6) | 111 (26.8) | 1.79 ± 0.84 |
| IO needle | 223 (53.9) | 88 (21.3) | 103 (24.9) | 1.71 ± 0.84 |
| Lab.support (Glucometer with strips, CBC) | 216 (52.2) | 99 (23.9) | 99 (23.9) | 1.72 ± 0.83 |
| Umbilical catheter | 186 (44.0) | 116 (28.0) | 116 (28.0) | 1.84 ± 0.83 |
| Iv cannulas different size | 174 (42.0) | 116 (28.0) | 124 (30.0) | 1.88 ± 0.84 |
| Cardboard to make splints | 174 (42.0) | 117 (28.3) | 123 (29.7) | 1.88 ± 0.84 |
| Adhesive tape and cotton | 165 (39.9) | 118 (28.5) | 131 (31.6) | 1.92 ± 0.84 |
| Thermometer | 173 (41.8) | 115 (27.8) | 126 (30.4) | 1.89 ± 0.84 |
| Manual BP cuff with a stethoscope | 170 (41.1) | 107 (25.8) | 137 (33.1) | 1.92 ± 0.86 |
| Electric suction pump with suction catheters | 162 (39.1) | 130 (31.4) | 122 (29.5) | 1.90 ± 0.82 |
| Nebulizer | 170 (41.1) | 120 (29.0) | 124 (30.0) | 1.89 ± 0.84 |
| Simple procedure (Foley catheter, dressing.) | 156 (37.7) | 118 (28.5) | 140 (33.8) | 1.96 ± 0.85 |
| Complex procedure (splints, pop.) | 156 (37.7) | 124 (30.0) | 134 (32.4) | 1.95 ± 0.84 |

BP = blood pressure. IO= intraossues. CBC = complete blood count. IV= intravenous.

### The knowledge of HCP toward essential equipment

The result of the study shows 241 HCPs (58.2%) had good knowledge, while 173 (41.8%) showed deficient understanding of essential equipment based mean score (</<u>></u>1.86). The evaluation of individual responses revealed distinct patterns in equipment recognition among healthcare providers (Table 8).

**Table 8:** Responses of healthcare providers regarding presence of management support, standardized guideline and protocols and essential drugs and laboratory support at a selected Tertiary Hospital in West Oromia, 2025.

| Variables | Responses |  |
| --- | --- | --- |
|  | Yes | No |
|  | N (%) | N (%) |
| Presence of management support | 183 (44.2) | 231 (55.8) |
| Presence of standardized guideline and protocols | 223 (53.9) | 191 (46.1) |
| Presence of essential drugs and lab. Support | 216 (52.2) | 198 (47.8) |

### Attitude of healthcare providers

The study results shows an attitudes of HCPs regarding pediatric emergency triage were 52.9% had good attitudes toward triage practices, while 47.1% had poor attitudes based mean score(</> 3.04) (Figure 6).

### Institutional-related factors

This study revealed that 55.8% of respondents, reported inadequate management and supervisory support for pediatric emergency triage. Additionally, 53.9% indicated that the absence of standardized guidelines and protocols had a significant effect on triage practice. Furthermore, 52.2% identified the lack of essential drugs and laboratory support as a critical obstacle to the proper implementation of pediatric emergency triage practice (Table 9)

**Table 9:** Institutional-related barriers to pediatric emergency triage regarding practice among healthcare providers in selected tertiary hospitals in West Oromia, Ethiopia, 2025 (n=414).

| Barriers | Categories | Frequency | Percentage |
| --- | --- | --- | --- |
| Workload | Yes | 246 | 59.4 |
|  | No | 168 | 40.6 |
| lack of time | Yes | 109 | 26.3 |
|  | No | 305 | 73.7 |
| Number of staff shortage | Yes | 58 | 14.0 |
|  | No | 356 | 86.0 |
| Lack of separate pediatric emergency rooms | Yes | 281 | 67.9 |
|  | No | 133 | 32.1 |
| Lack of three color coded | Yes | 77 | 18.6 |
|  | No | 337 | 81.4 |
| Lack of interest at triage | Yes | 128 | 30.9 |
|  | No | 286 | 69.1 |
| Unfamiliarity with protocols, medications or indications for practice triage | Yes | 242 | 58.5 |
|  | No | 172 | 41.5 |
| Lack of internet access | Yes | 78 | 18.8 |
|  | No | 336 | 81.2 |
| Lack of seniors staff | Yes | 273 | 65.9 |
|  | No | 141 | 34.1 |
| Language barriers between patient and healthcare providers | Yes | 105 | 25.4 |
|  | No | 309 | 74.6 |
| Insufficient availability of training for healthcare providers | Yes | 248 | 59.9 |
|  | No | 166 | 40.1 |
| Healthcare providers cultural or religious believes | Yes | 44 | 10.6 |
|  | No | 370 | 89.4 |

The study identified several key barriers affecting practice of pediatric emergency triage. Three predominant challenges emerged: overwhelming clinical workloads (affecting 59.4% of providers), absence of specialized pediatric emergency spaces (67.9%), and critical shortages of experienced medical personnel (65.9%). Compounding these issues, nearly 60% of healthcare workers cited inadequate training programs (59.9%) and insufficient familiarity with essential triage protocols and pediatric medications (58.5%), revealing substantial educational and operational deficiencies. Most of the participants reported that the challenges of healthcare providers were workload 246 (59.4%) and the lack of senior staff (65.9%), which impacted patient triage, categorization, and allocation. However, addressing these barriers challenges through effective resource allocation, comprehensive training programs, and improved communication strategies might be enhanced to practice triage efficiency and improve pediatric emergency triage outcomes (Table 10).

**Table 10:**
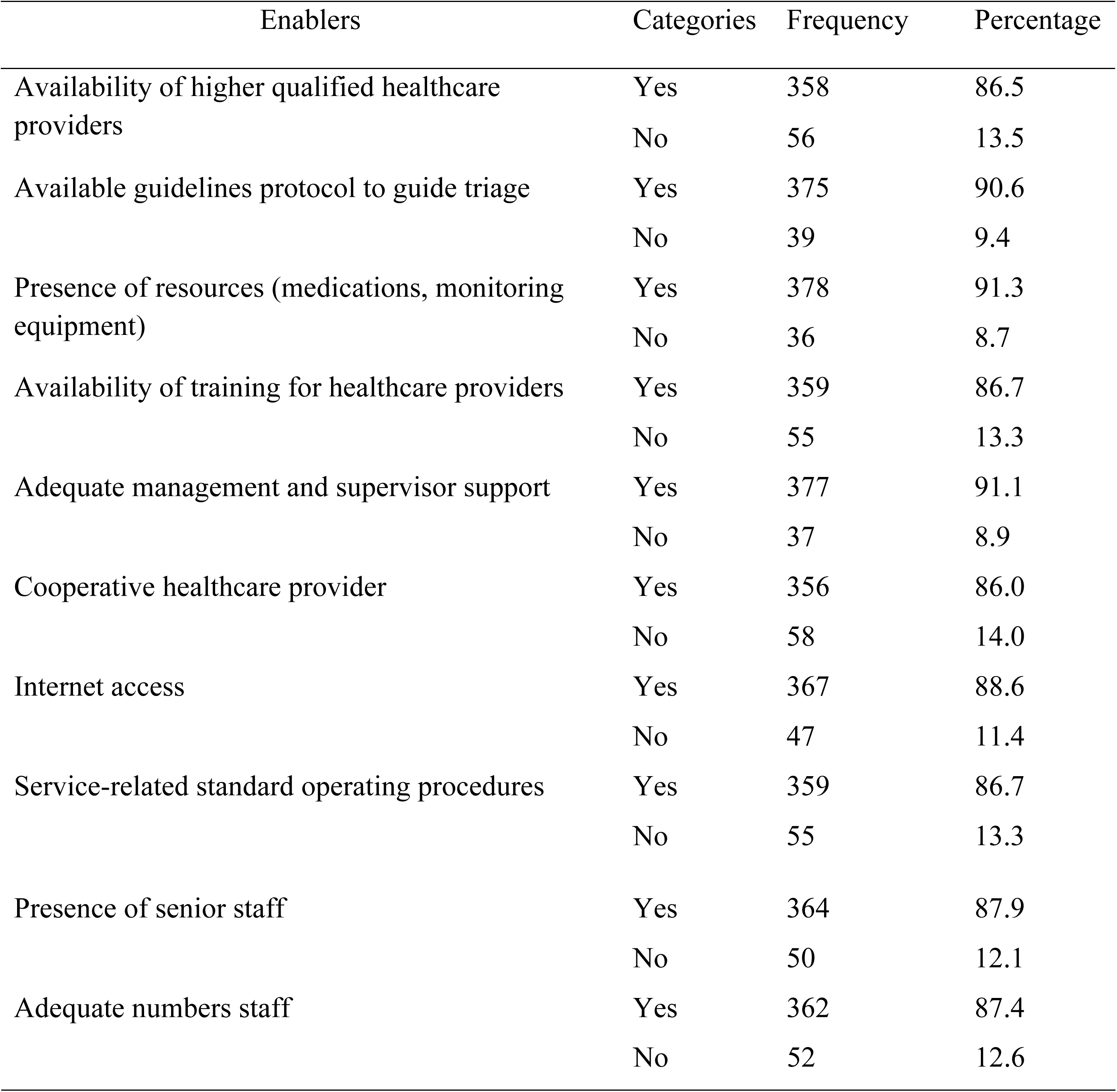
Institutional-related enablers to pediatric emergency triage concerning practice among healthcare providers in selected tertiary hospitals in West Oromia, Ethiopia, 2025 (n=414).

### Enablers of emergency triage practice

The study result shows the demonstrated strong institutional capacity for pediatric emergency triage across multiple domains. Human resources were well-established, with 87.9% of facilities reporting available senior staff, 87.4% maintaining adequate staffing levels, and 86.5% confirming the presence of highly qualified healthcare providers (Table 11).

**Table 11:**
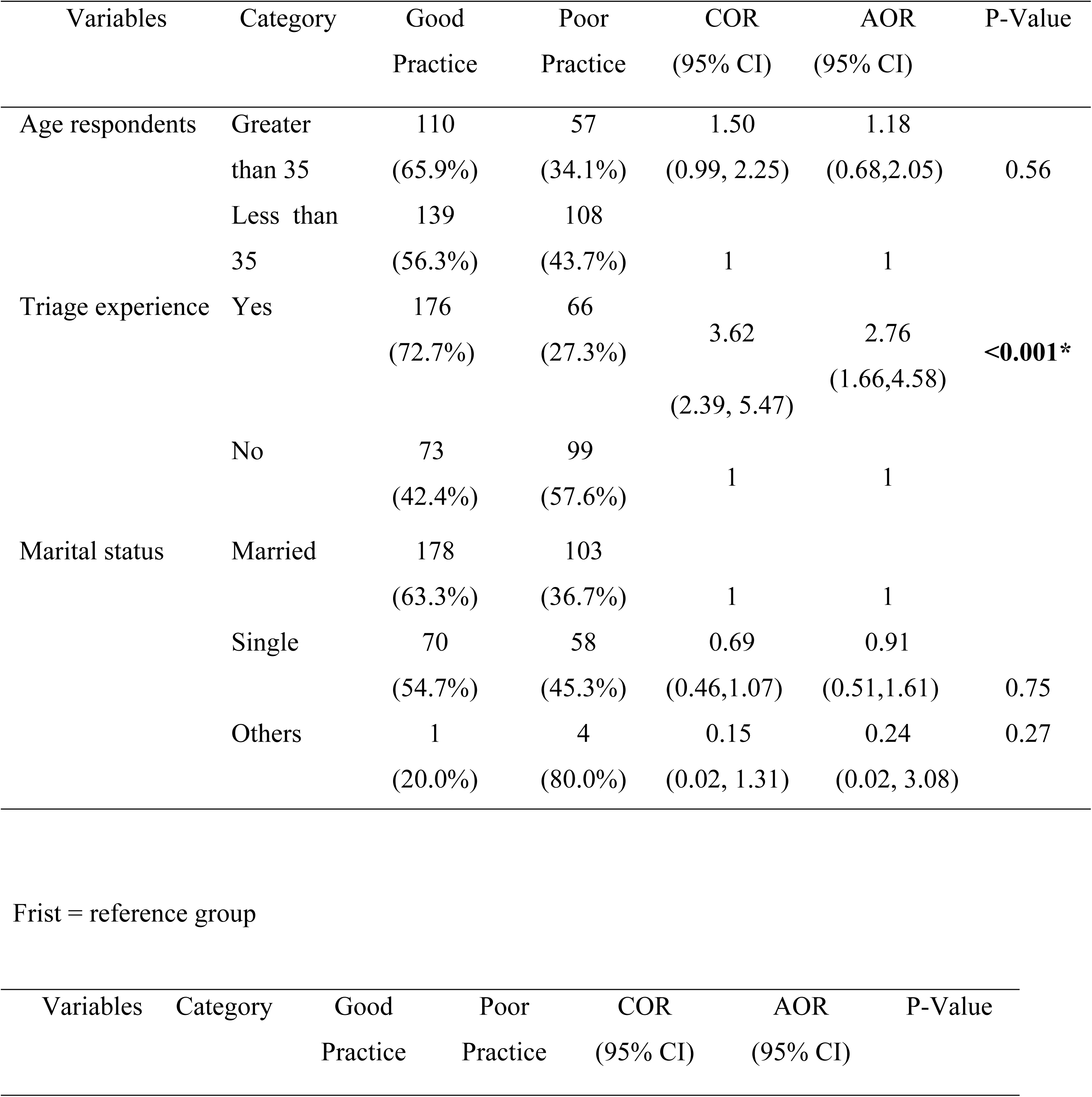
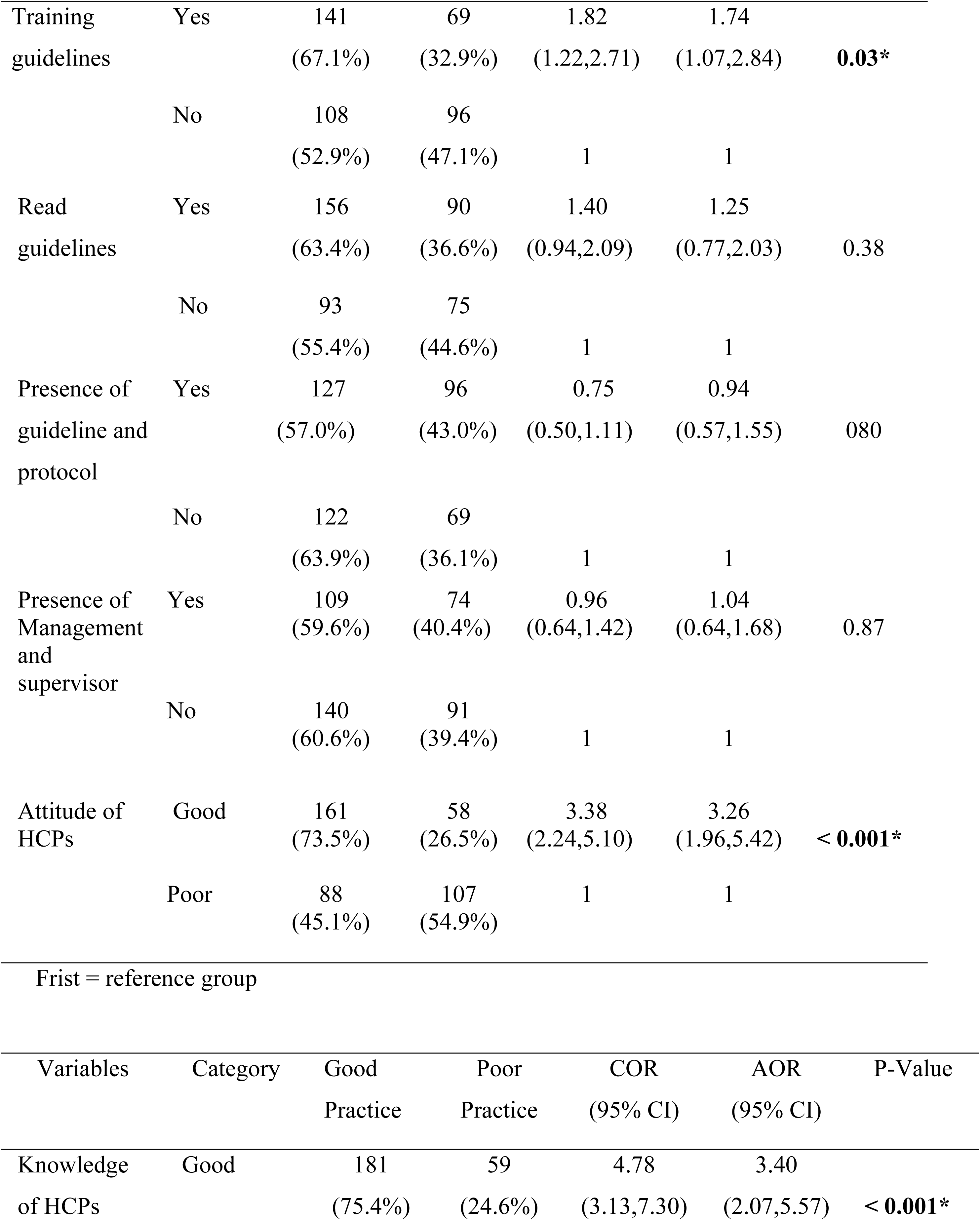
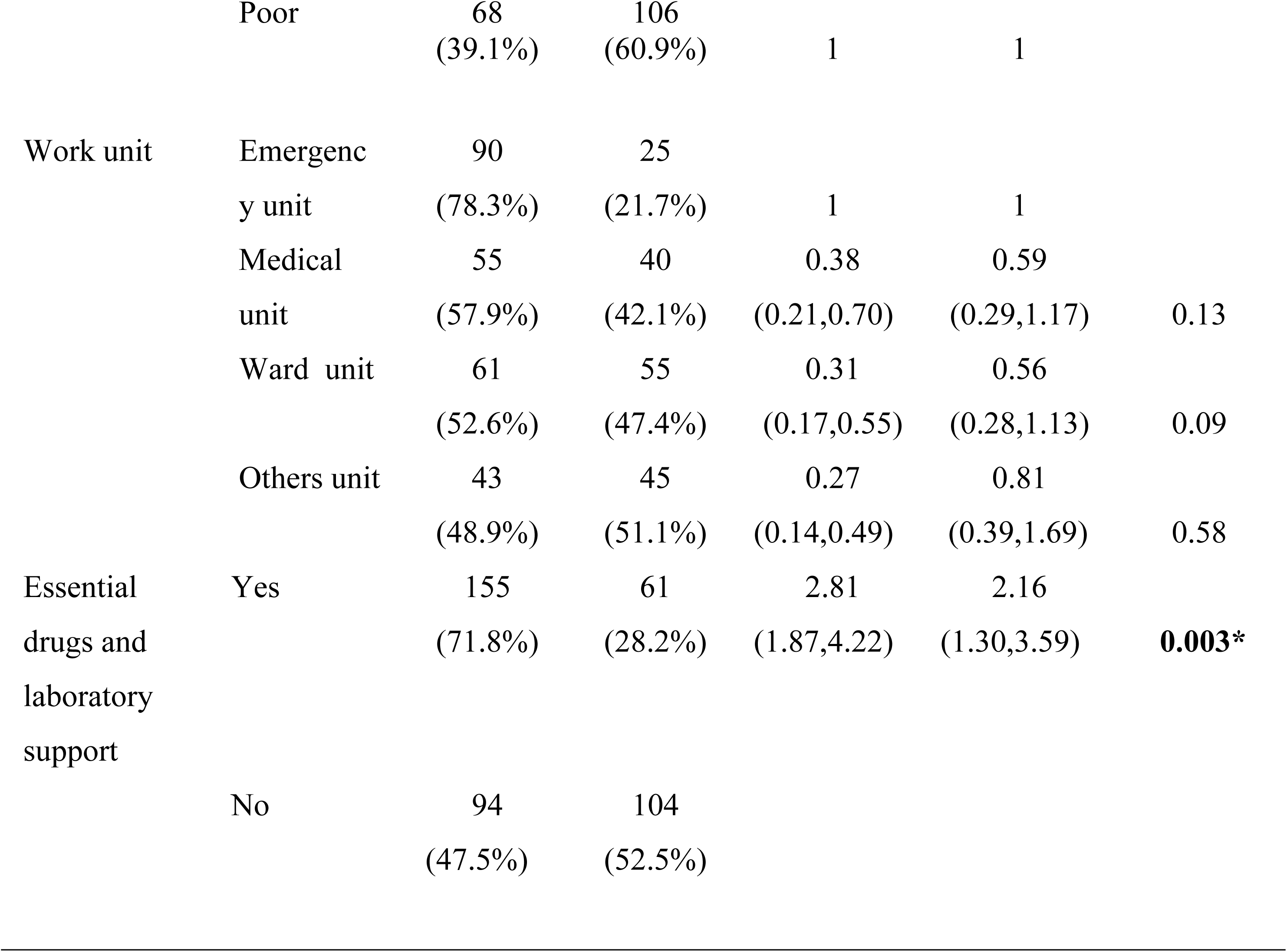
Bivariate and multivariate binary logistic regression analysis of practice and associated factors among healthcare providers working in tertiary hospitals, west Oromia, Ethiopia, 2025 (n=414).

### Factors associated with pediatric emergency triage practice

In bivariate analysis, 11 were identified as candidate variables for multivariable logistic regression based on a p-value < 0.25 significance level. These variable included triage experience, training guidelines, age, marital status, work unit, read guidelines, knowledge of HCPs, presence of management and supervisor, presence of standardized guidelines and protocols, essential drugs and laboratory support, and the attitude of healthcare providers.

After adjusting for other covariates, the multivariate binary logistic regression model revealed that five variables were significantly associated with CF at p ≤ 0.05, with a 95% confidence interval. These five variables were Knowledge of HCPs, presence of essential drugs and laboratory support, triage experience, training guidelines, and the attitude of healthcare providers. The logistic regression model was statistically significant (X^2^ = 139.082; df = 19; p < 0.001). The model also showed that 39.0 % (Nagelkerke R^2^) of the variance in the practice score among healthcare providers was explained by the combined effects of those independent variables, and correctly classified 75.1% of cases overall.

Accordingly, after adjusting for other covariates, the odds to demonstrate good triage practices were nearly twice as likely [AOR = 1.74, 95% CI: (1.07 - 2.84); P = 0.03] among those who got training guideline than those who did not get training guideline. Additionally, healthcare providers having good knowledge on triage practice were three times more likely to have good triage practice [AOR = 3.40, 95% CI: (2.07 - 5.57); P = < 0.001] compared to their counterparts. Morever, healthcare providers who exhibited positive attitudes toward triaging practices were 3.26 times more likely to engage in good triage practices [AOR = 3.26; 95% CI: (1.96 - 5.42); P = < 0.001] compared to those with poor attitudes (Table 13).

The participants with prior triage experience were nearly three times more likely to demonstrate good triage practices (AOR = 2.76; 95% CI: 1.66–4.58; P < 0.001) compared to those without triage experience. Finally, the study revealed that, healthcare providers with access to essential drugs and equipment were 2.16 more likely to adopt good triage practices (AOR = 2.16; 95% CI: (1.30 - 3.59); P = 0.003) compared to those lacking these triage resources (Table 13).

## DISCUSSION

The purpose of this study is to investigate the practice of pediatric emergency triage and its associated factors among healthcare providers working at tertiary hospitals in West Oromia. Several key factors influence the practice of pediatric emergency triage, including healthcare providers’ knowledge of ETAT, the availability of essential drugs and laboratory support, triage experience, training on ETAT guidelines, and their attitudes toward triage.

The study revealed that 60.1 % (95 % CI (55.2%, 64.9%)) of the respondents had good practice of pediatric emergency triage, indicating that nearly three in five healthcare providers reported to have a good practice of pediatric emergency triage. In the present study, the prevalence of good practice in pediatric emergency triage was higher than that reported in studies conducted in Addis Ababa, Limpopo, Rwanda and Tanzania, where only 52.9%, 37%, 52.1% and 52% of healthcare providers, respectively, demonstrated good practice in pediatric emergency triage (11,12,16,17).

The difference might be due to variations in sample size and the inclusion of only nursing professionals. Variations in sample composition likely contributed. For instance, the Addis Ababa study included only 197 nurses (16), while our study incorporated multidisciplinary healthcare providers among whom doctors may possess more field experience and longer tenure factors associated with triage practice, This suggests that professional diversity in our sample may have elevated overall triage practice performance (18). Also, this discrepancy may be attributed to the incremental implementation of triage protocols, time of study which suggests potential improvements in systems and practices of triage at our study site (19).

The prevalence of good pediatric emergency triage practice in this study was lower compared to findings from prior research conducted in Keniya, Jordan and Indonesia where 65.4%,84.8%,79.1% participants demonstrated good practice in pediatric emergency triage respectively(20–22). This discrepancy may be attributed to differences in study setting, variability in professional training and experience, disparities in resource availability, as well as sample size variations across studies. Additionally, healthcare providers working during the malaria were greatly expanded in west Oromia and are exposed to many patients difficult to triage in a short time, making them more prone to practice.

The study finding shows a statistically significant positive association between training guideline and practice of paediatric emergency triage. Healthcare providers who received training on guidelines were nearly twice as likely to demonstrate good practices compared to those who did not receive training. This finding similar with studies conducted in KwaZulu-Natal, Gondar, and Wolaita (13,23,24). This might be attributed to received training enhances knowledge and builds practical skills of HCP on practice of pediatrics triage, thereby enabling providers to accurately assess and prioritize emergency cases (13).

This might be attributed received training in triage enhances the efficiency and effectiveness of triage practice among healthcare providers (25). Also as their confidence grows through formal education, they become better equipped to perform triage accurately and efficiently (26), Additionally, implementing ongoing training programs for nurses specializing in triage ensures continuous skill development and improves overall practice patient assessment and management (23,27,28). This study related seek to establish a robust training framework designed to enhance the accuracy of pediatric emergency triage, decision-making in patient prioritization and effective triage plays a crucial role in ensuring patient safety (29,30).

Additionally, healthcare providers with a good knowledge on triage practice were more likely to have good practice compared to their counterparts with poor knowledge. This result is consistent with study from Indonesia (20,26). This might be attributed to those healthcare providers with good knowledge are capable of making swift, informed decisions that ensure patients receive the appropriate level of care urgently (8,10,31). Also having adequate knowledge on triage leads to greater confidence and consistency in following established guidelines, reducing errors in patient triage (32,33). According to the knowledge-attitude-behavior (KAB) model, it serves as a framework for understanding how information is processed and translated into action. While good knowledge acquisition is crucial, ensuring that it leads to practice consideration of precursor influencing factors emergency triage (34,35).

This study finding shows a statistically significant positive association between triage experience and the level of practice among healthcare providers. The study also revealed that participants with triage experience were more likely to demonstrate good practices compared to those who did not have triage experience. This study finding was supported by the studies conducted in Hawasa and Limpopo (10,22). This might be due to having triage experience helps in developing the critical decision-making skills necessary for effective triage practice (36). Also having prior experience in triage increases triage exposure, which is key factor in achieving higher proficiency in emergency care and triage (7,22,32). Similarly, according to Benner’s five-stage theory of skill acquisition, skill development is primarily influenced by years of triage experience. However, the demand for proficient and expert healthcare in triage remains high. The ability to integrate diverse types of knowledge enhances its applicability across various clinical situations, ultimately improving triage practice outcomes (32,37).

Moreover, the study revealed healthcare providers who exhibited positive attitudes toward triaging practices were more likely to engage in good triage practices compared to those with poor attitudes. This finding is similar with studies conducted in Indonesia, Gaza Strip, and Hawasa (10,32,38). This finding supports that having good attitudes play great role in shaping clinical and triage performance (39). Furthermore, healthcare providers who have positive attitude towards pediatric emergency triage tend to adhere more closely to guidelines and protocols (40). This finding is supported by study conducted in Ghana that reported most healthcare providers had a good attitude of their knowledge and understanding of triage in emergency departments (41). The study finding highlighted the need for regular workshops and in-service training to enhance HCP skills.

Finally, the study’s results have also shown that availability of essential drugs and equipment had a significant positive association with practice of pediatric emergency triage. This result aligns with previous research conducted in Hawasa, Indonesia Amman, Jordan and Saudi Arabia (10,14,20,22,42). Also, this study finding was supported by similar study results in developing countries like Ethiopia where economic factors highly challenges practice of emergency triage (43).

A possible explanation is that the presence of essential drugs and equipment significantly influences pediatric emergency triage practices. The availability of these critical resources plays a vital role in improving pediatric emergency triage practice (44). Enhancements in this area could lead to better patient outcomes, increased operational efficiency, and improved working conditions for healthcare providers (18,45). Moreover, ensuring strong resource accessibility in pediatric emergency triage settings is fundamental to optimizing care delivery and enabling effective responses to critical medical situations (42,46). Ensuring optimal adherence to PET protocols requires a multifaceted approach, including improved training, standardized guidelines, and enhanced resource allocation (47,48). Addressing these challenges will be contribute to more reliable emergency care and better pediatric emergency patient outcomes (40).

## Conclusion

The study revealed that more than three fifth of healthcare providers reported having a good level of triage practice. This finding indicates that key predictors influencing triage practice include the presence of essential drugs and laboratory support, knowledge of healthcare providers, triage experience, training of guideline, and attitudes of healthcare providers which may adversely impact practice of pediatric emergency triage and healthcare outcomes. However, efficient pediatric emergency triage practice relies on essential medications and laboratory support, as well as healthcare providers’ knowledge, triage experience, attitudes, and guideline-based training. These factors enhance decision-making and adherence to protocols, ensuring consistency in triage practices. Therefore, to improve accuracy and efficiency, targeted interventions and evidence-based strategies are essential for addressing gaps and optimizing patient care and triage practice.

This study indicates that hospital administrators should pay greater attention to the well-being of healthcare providers and prepare training programs that help healthcare provider increase triage practice strategies. They should promote teamwork and support among healthcare providers, friends, nursing managers, and other healthcare providers. Additionally, administrators should assess triage experience when rotating healthcare providers to different work units, particularly pediatric emergency as these factors influence the risk associated with triage practices and take measures to prevent it. Also the healthcare providers should adopt strategies such as getting training on guideline, developing individuals’ knowledge skills, and improving their attitude towards pediatrics emergency triage.

Furthermore, EMA and ENA should serve as an advocate for well-being and satisfaction of healthcare providers’, ensuring that doctors and nurses are more aware practice of pediatric emergency triage and their impacts on patients outcomes, and the health care system. NGOs should also provide the financial and material support needed for training and targeted activities to more increase triage practice.

## Declarations

### Ethics approval and consent to participate

Ethical clearance for this study was obtained from the Institutional Review Board (IRB) of the Institute of Health Sciences Research at Wallaga University (Ref. No: IHSREC103/104/2025). Additionally, a letter of support was secured from the Nursing and Midwifery Academic and Service Director Office (REF: NMASDO/176/2017) for submission to each hospital, followed by official permission from the hospital administrations to conduct the study. The medical director of each hospital was formally requested to facilitate cooperation during data collection. The head nurse of each unit was briefed on the study’s objectives and subsequently informed the nursing staff. All participants were clearly informed about the purpose of the study, their voluntary participation, and their right to withdraw at any time without consequence. The study strictly adhered to the principles of the Helsinki Declaration. Participants were assured of confidentiality, with no personal identifiers linked to their responses. Written informed consent was obtained from all respondents prior to data collection.

### Consent for Publication

Not Applicable

### Availability of data and materials

Data will be available from corresponding author upon reasonable request

## Authors’ contributions

All authors made substantial contributions to conception and design, acquisition of data, or analysis and interpretation of data; took part in drafting the article or revising it critically for important intellectual content; gave final approval of the version to be published; and agree to be accountable for all aspects of the work.

## Data Availability

All relevant data are within the manuscript and its Supporting Information files

## Acknowledgements

We want to thank West Oromia Tertiary hospital administrators, data collectors, supervisors, and respondents.

## Acronyms/Abbreviation

ABCDE: Airway, Breath, Circulation, Dehydration, and Exposure
AURH: Ambo University Referral Hospital
DDUCSH: Dambi Dollo University Comprehensive Specialized Hospital ED Emergency Department
ETAT: Emergency triage assessment and treatment
HCP: Healthcare Providers
IMNCI: Integrated management of childhood illness
JUMC: Jimma University Medical Center
MKCSH: Mettu Karl Comprehensive Specialized Hospital
MOB: Malnutrition, oedema and Burns
NCSH: Nekemte Comprehensive Specialized Hospital
OPD: Outpatient department
PAT: Paediatric assessment Triage
PED: Paediatric emergency department
SATS: South Africa triage scale
WHO: World Health Organization
WU: Wallaga University
WUCSH: Wallaga University Comprehensive Specialized Hospital

---

**Figure 1:**
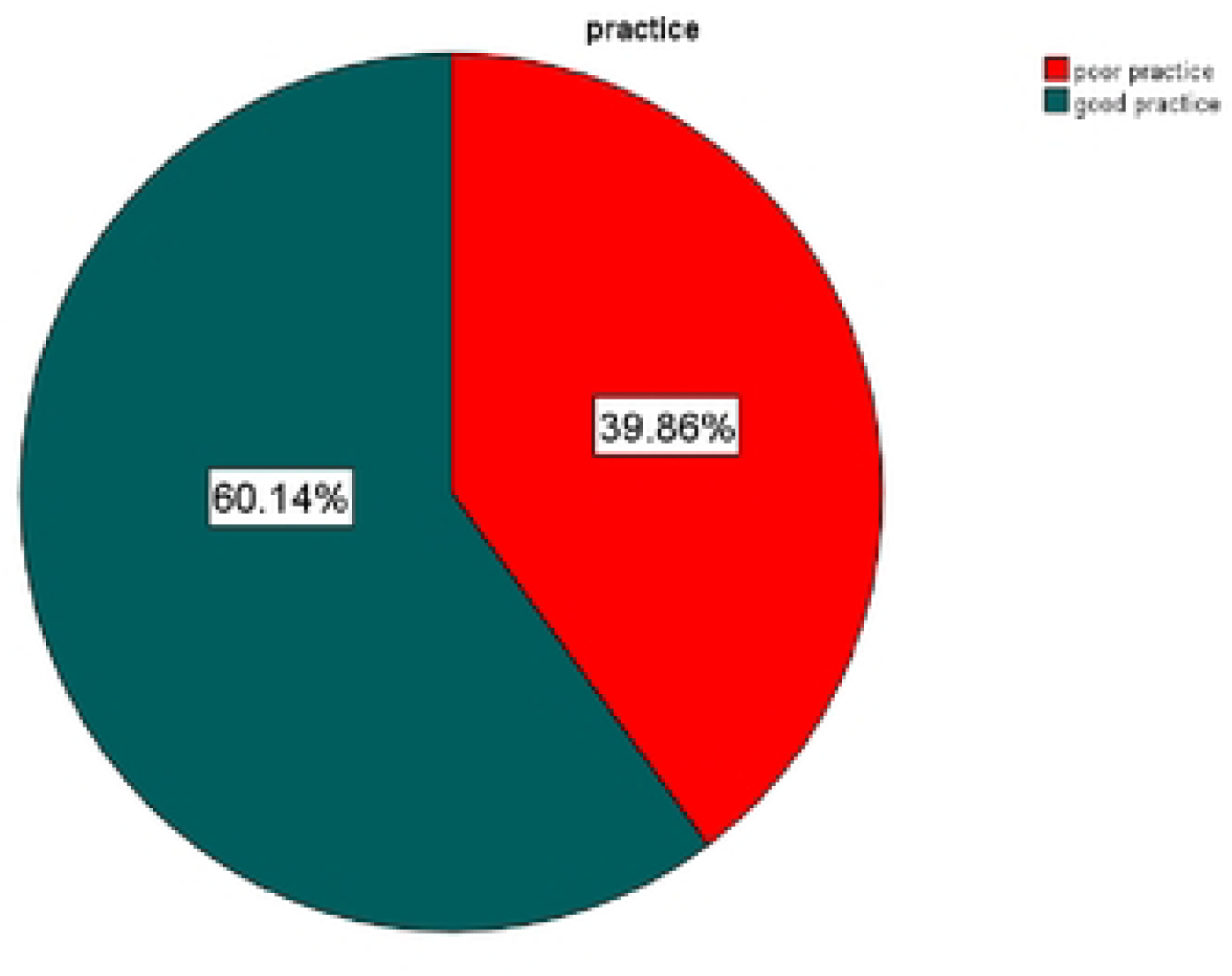
Level of practice in pediatric emergency triage among healthcare providers working in tertiary hospitals, west Oromia, Ethiopia, and 2025 (n=414).

**Figure 2:**
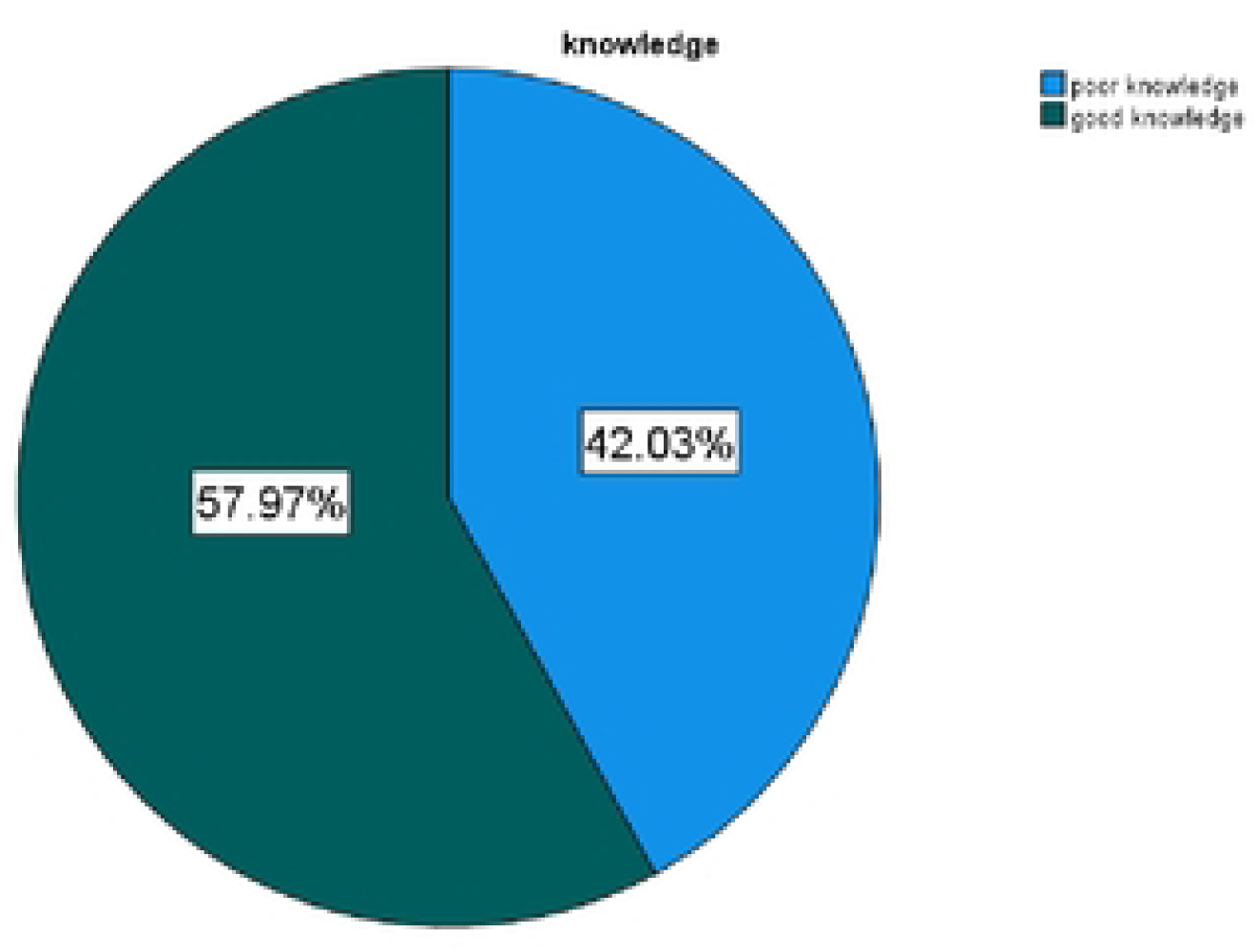
Overall knowledge levels of pediatric emergency triage among healthcare providers working in a tertiary hospital in west Oromia, Ethiopia, 2025, (n=414).

**Figure 3:**
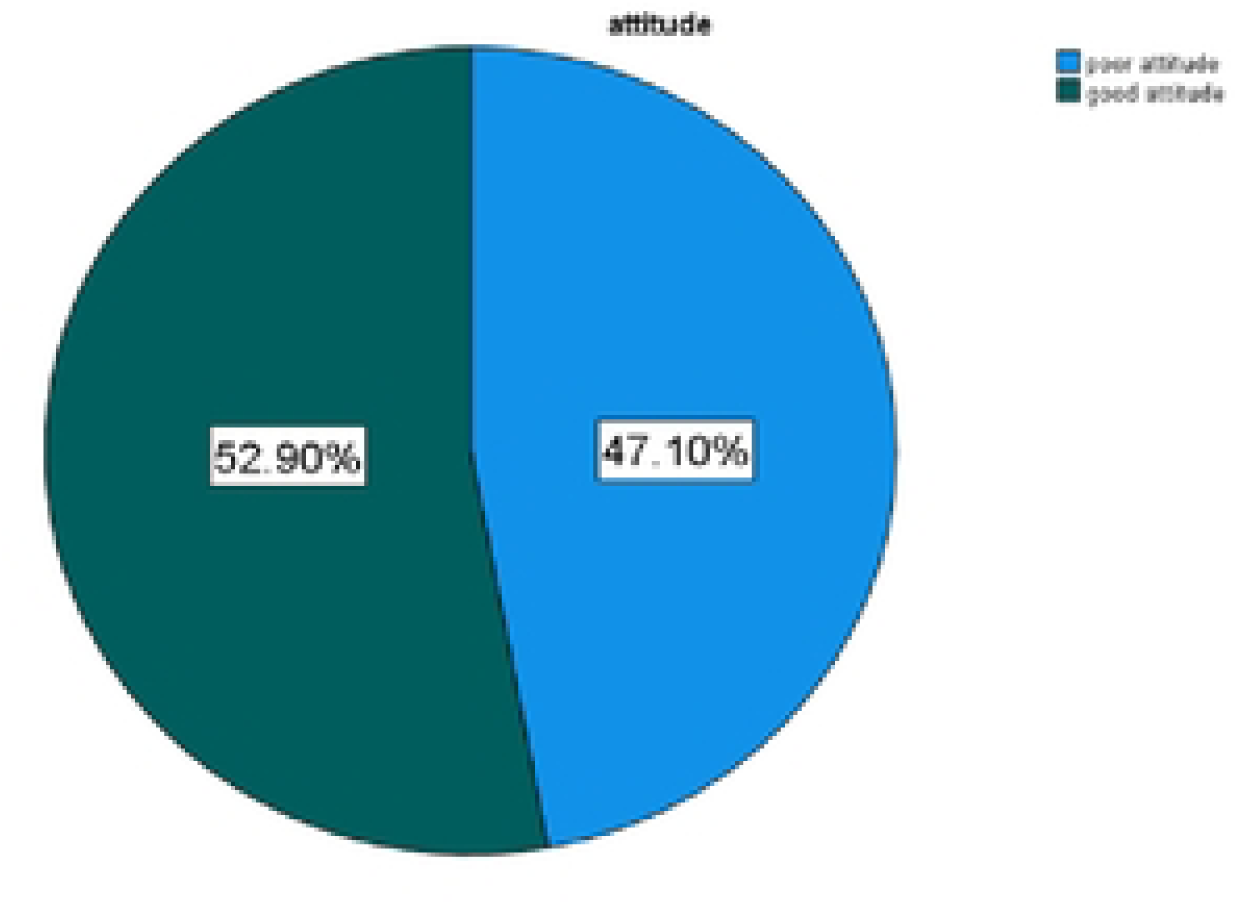
Attitude levels toward pediatric emergency triage among healthcare providers working in a tertiary hospital in West Oromia, Ethiopia, 2025 (n = 414).

